# Socio-demographic differences in the risk of suicides in children and young people: a population level linked study in England, 2011 to 2022

**DOI:** 10.1101/2025.05.01.25326827

**Authors:** Emma Sharland, Rachel Mullis, Emyr John, Isobel Ward, Cathryn Rodway, Daniel Ayoubkhani, Vahé Nafilyan

**Affiliations:** Health Research Group, Office for National Statistics, Newport, UK NP10 8XG; Population Health Sciences, Bristol Medical School, University of Bristol, Bristol, UK, BS12NT; The National Institute for Health and Care Research Applied Research Collaboration West (NIHR ARC West) at University Hospitals Bristol and Weston NHS Foundation Trust, UK; National Confidential Inquiry into Suicide and Safety in Mental Health (NCISH), Centre for Mental Health and Safety, Division of Psychology and Mental Health, Faculty of Biology, Medicine and Health, University of Manchester, Manchester M13 9PL

## Abstract

**Background:** Suicide is one of the leading causes of death in children and young people (CYP) globally. Over the past decade there has been a steady increase in the number of suicide deaths in CYP in the UK. This study aims to identify socio-demographic differences in the risk of suicide in CYP.

**Methods:** Using linked 2011 Census and death registrations data, we created a cohort of 7,747,345 CYP aged 10 to 17 years in England. We estimated adjusted incidence rate ratios (IRRs) using generalised linear models with a Poisson link function, to identify socio-demographic characteristics associated with death by suicide in CYP.

**Results:** The rate of suicide was highest in males compared to females (IRR = 0.6, 95% CI = 0.5 – 0.7) and in CYP of Mixed or Multiple ethnic groups (IRR = 1.5, 95% CI = 1.1 – 2.0) compared to those who were White. We also found an increased risk of suicide where the HRP’s highest level of education was a degree-level or above qualification (IRR = 1.5, 95% CI = 1.1 – 1.9) compared to households where the HRP had no qualifications.

**Conclusion:** Our population-level findings suggest CYP living in households with better educated parents may be one of the key groups most at risk of dying by suicide in England. Whilst our findings support some similar risk factors in CYP as have been found for adults, the relationship between measures of socio-economic status and risk of suicide appears to be more complex in CYP.

**Key Messages:** *What is already known on this topic:*

- There is limited evidence using population-level data in England for identifying inequalities in the risk of suicide in children and young people (CYP).
- Available evidence indicates higher rates of suicide in CYP who are: male, of White or Mixed or Multiple ethnic background, have poor mental health and a history of self-harm.
- Most research on the risk factors for suicide in CYP focuses on educational and familial issues such as marriage breakdowns and bereavements.

*What this study adds:*

- This study is the examines the socio-demographic differences in the risk of suicide in CYP aged 10 to 17 years old using administrative and census data with near-whole population coverage in England.
- We found similar inequalities for suicide in CYP that have been previously reported in adults. However, in contrast to previous research, we also found a higher risk of suicide in CYP where the head of household had a higher level of education.

*How this study might affect research, practice or policy:*

- Our findings highlight the key groups of CYP who are at a higher risk of suicide in England.
- Our results may be used in the development of future public health policies or interventions targeting these high-risk groups to reduce the risk of suicide in CYP.

## Introduction

Suicide is a major public health concern and among children and young people (CYP) aged 10-to 24-years old. In 2023, suicide was the leading cause of death in England for 5-to 34-year-olds (14.5% of deaths in 5-to 19-year-olds; 23.1% in 20-to 34-year-olds)^1^. In 2024, the World Health Organization (WHO) declared suicide the third leading cause of death among 15-to 29-year-olds globally^2^. While CYP suicide rates are relatively low compared with older age groups, rates have been increasing over the last decade, particularly in females^3^, for example, in England for 15-to 19-year-olds, the rate of suicide in 2013 was 3.8 per 100,000, increasing to 5.2 per 100,000 in 2023^3^. The 2023 Suicide Prevention Strategy for England^4^ identifies CYP as a priority group to provide tailored and targeted support and calls for improved evidence to better understand their experience.

Mental ill-health has also risen in recent years in CYP with around 1 in 5 reporting a probable mental disorder in 2023^5^ and reports indicating an 84% increase in referrals to CYP’s mental health services in England between 2018-2019 and 2021-2022^6^. The development of suicide risk is complex and often involves a combination of biological, psychological, clinical, social, and environmental risk factors^7^. CYP can be particularly vulnerable to mental health stressors magnifying these factors due to CYP still developing neurocognitively and bring unable to assess long-term risks of their actions^8^. Suicide prevention strategies are complex to deliver, so knowing the groups of CYP to target for these interventions is important.

Most studies identifying risk factors for suicide in CYP focus on health (including self-harm, other mental health conditions, prescribed medications), educational and familial characteristics^9–11^, with limited evidence from England on the socio-demographic differences in risk. Research has identified higher rates of suicides in children who are male, of White or Mixed or Multiple ethnic background and in those who have previously self-harmed, and those living in deprived areas^9,11,12^. The literature on SES as a risk factor for suicide in adults has linked lower skilled occupations and lower levels of education with a higher risk of suicide^13,14^. There is little research investigating the link between SES and the risk of suicide in CYP, whose SES will likely be driven by characteristics of adult household members. Some evidence does support CYP from low SES background being at elevated risk of suicide, as CYP with low parental levels of education found to be related to an increased risk of suicidal ideation in young people^15^.

Few England-based studies have investigated socio-demographic risk factors for suicide in CYP using nationally representative samples while also adjusting for potential confounding differences between at-risk groups. Therefore, the aim of this study is to investigate the association between socio-demographic characteristics and the risk of suicide in CYP using administrative datasets linked with the 2011 Census, with near-whole population coverage in England.

## Methods

### Study data and population

We linked Census 2011 and death registrations by NHS number for usual residents in England. NHS number was obtained by linking the 2011 Census to the 2011-2013 NHS Patient registers (93% linkage rate)^16^. Individuals were included if they were aged between 10 and 17 years on Census Day (27 March 2011) or from their 10^th^ birthday if they were aged under 10 on Census Day. Individuals were followed until the earliest of; end of study period, their 18^th^ birthday or death. Ethical approval was obtained from the National Statistician’s Data Ethics Advisory Committee (NSDEC(22(17)).

### Exposures

Individual exposure variables explored in this analysis were sex, ethnic group, religion, disability status, main language, region of residence. Household exposures were National Statistics Socio-Economic Classification (NS-SEC), highest level of qualification and religion, of the household reference person (HRP) (Supplementary table 1). Due to the age of the CYP in this study, it is likely that the exposures included in this study have been reported by proxy for the CYP by the main Census respondent in the household.

### Outcomes

Our primary outcome was death due to suicide between 28 March 2011 to 31 December 2022. For comparison we also looked at a secondary outcome of non-suicide death (any underlying cause of death that was not classified as suicide). For suicides, the median registration delay for deaths registered in 2023 was 199 days in England, while the median delay for all deaths was 7 days in 2022^3,17^. Therefore, to reduce any potential bias arising from delays in death registration, we used deaths registered by 31 December 2023.

Suicide was defined as death from intentional self-harm for persons aged 10□years and over (ICD-10 codes X60-X84), and death caused by injury or poisoning where the intent was undetermined for those aged 15□years and over (ICD-10 codes Y10-Y34)^18–20^ (Supplementary tables 2 and 3).

### Statistical Analyses

Descriptive statistics of population characteristics were calculated stratified by all outcomes (death by suicide and non-suicide deaths) along with rates of death per 100,000 person-years during the study period. To model the association between socio-demographic characteristics and the risk of suicide, we fitted generalized linear models with a Poisson link function, with death by suicide being the outcome of interest. Age was calculated on Census modelled as a natural spline with boundary knots at the 1st and 99th percentiles and one internal knot at the median of the age distribution. An offset was added to the model to account for the different time-at-risk periods between individuals (transformed with a natural logarithm). Time at risk was defined as days from 2011 Census Day (or date of 10th birthday) to date of death (by any cause), date of 18^th^ birthday or end of study (31 December 2022), whichever was earlier.

For each exposure, to estimate the difference in the rate of suicide, we fitted models minimally adjusted for age and sex. To assess the independent associations between our primary exposures and risk of suicide, we then fitted models adjusted for a wider range of covariates. For all exposures (except disability) in the adjusted model, we controlled for age, sex, ethnic group, main language, carer status, disability status, NS-SEC^21^, qualifications, and region of residence. For disability status, we controlled for age, sex, ethnic group, main language, NS-SEC, qualifications, and region of residence, we did not control for carer status due to the low likelihood of CYP reporting having a disability and also being carers.

Statistical significance was determined at the 5% level, and all reported confidence intervals are at the 95% level. Data preparation was conducted using Sparklyr version 3.3.0, and statistical analysis was performed using R version 4.4.

## Results

### Descriptive Statistics

Our study population consisted of 7,747,345 CYP aged between 10 and 17 years during the study period (27 March 2011 to 31 December 2022; see Supplementary table 4). The linkage rate for the final study population was 96.4%. In our cohort, 41.5% of the sample were aged 10 to 17 on Census Day and 58.5% turned 10 years old after Census Day but died before the end of study. The mean follow-up was 5.4 years. The study population was 51.1% male and 80.8% were from a White ethnic background (Table 1). There 710 suicides over the study period, predominantly in males (63.4%) and in the white ethnic group (83.8%; Table 1).

**Table 1:**
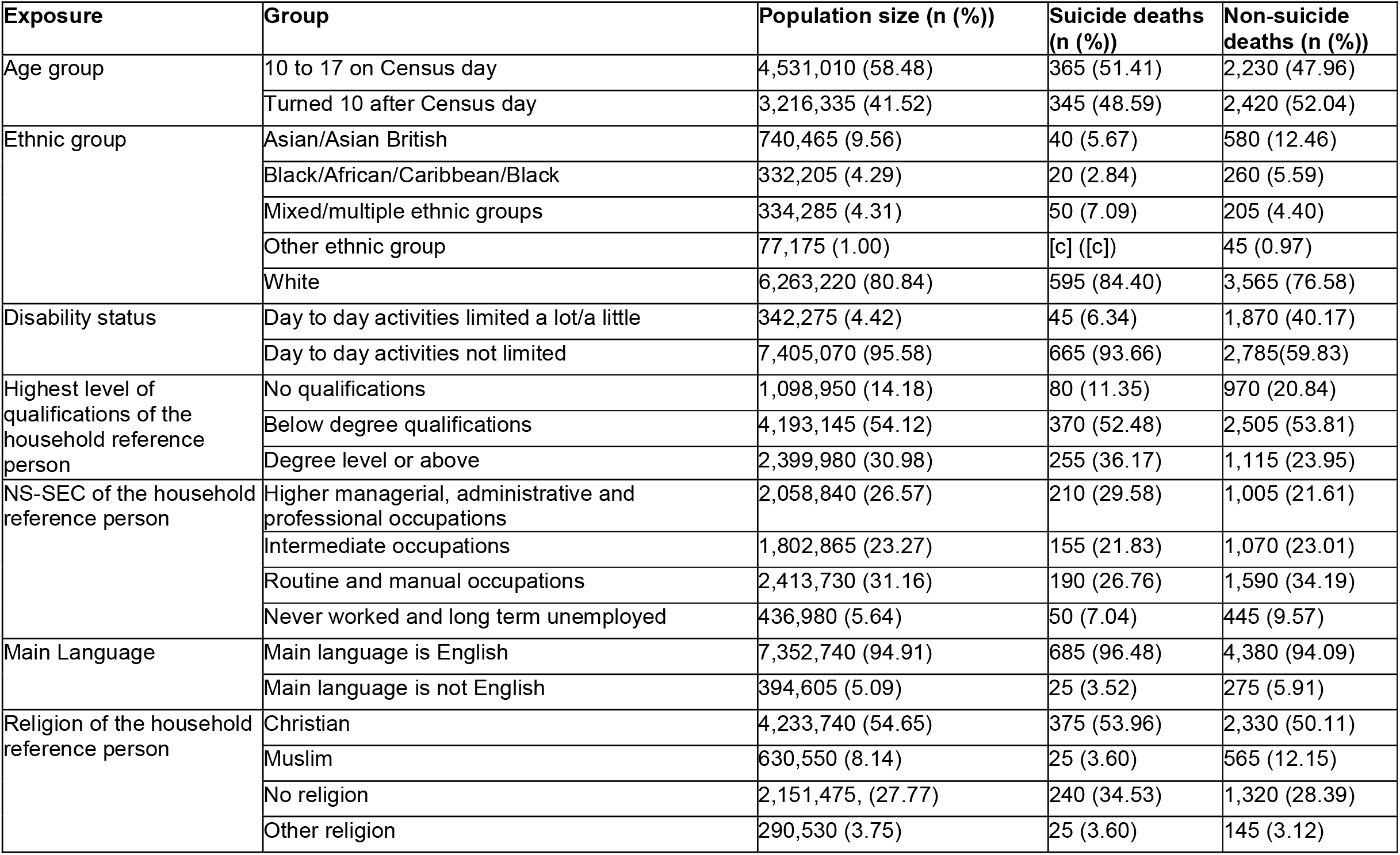
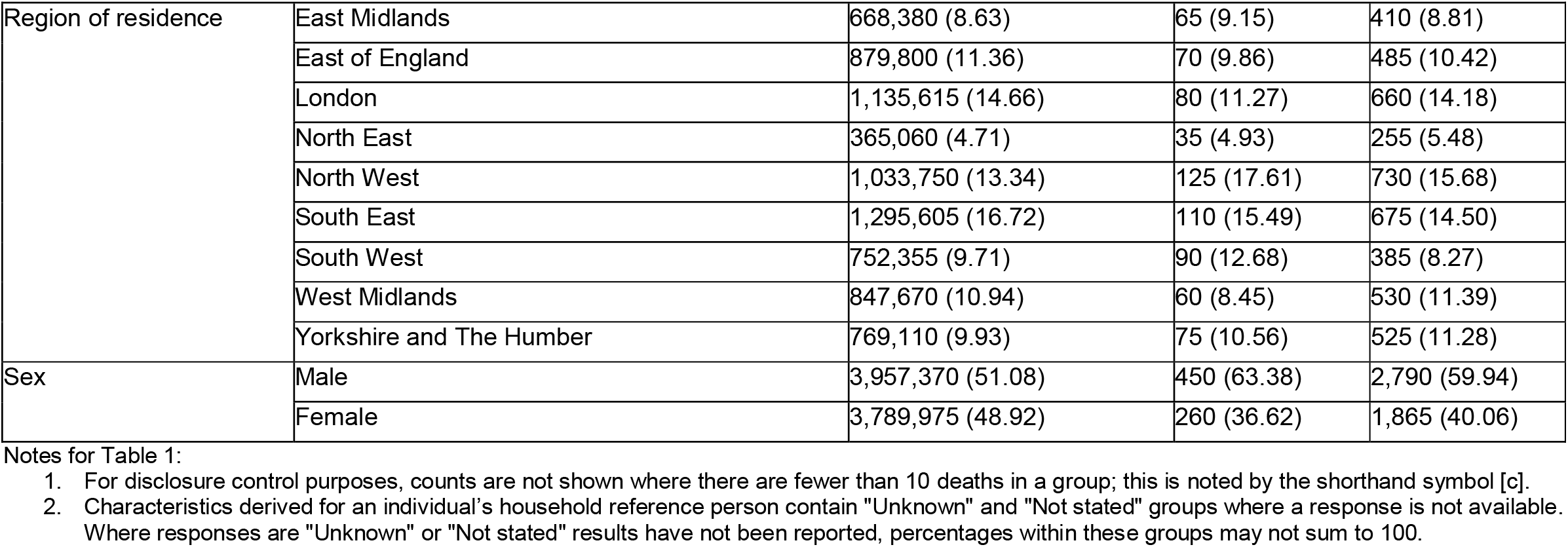
Descriptive statistics of the cohort.

| **Exposure** | **Group** | **Population size (n (%))** | **Suicide deaths (n (%))** | **Non-suicide deaths (n (%))** |
| --- | --- | --- | --- | --- |
| Age group | 10 to 17 on Census day | 4,531,010 (58.48) | 365 (51.41) | 2,230 (47.96) |
|  | Turned 10 after Census day | 3,216,335 (41.52) | 345 (48.59) | 2,420 (52.04) |
| Ethnic group | Asian/Asian British | 740,465 (9.56) | 40 (5.67) | 580 (12.46) |
|  | Black/African/Caribbean/Black | 332,205 (4.29) | 20 (2.84) | 260 (5.59) |
|  | Mixed/multiple ethnic groups | 334,285 (4.31) | 50 (7.09) | 205 (4.40) |
|  | Other ethnic group | 77,175 (1.00) | [c] ([c]) | 45 (0.97) |
|  | White | 6,263,220 (80.84) | 595 (84.40) | 3,565 (76.58) |
| Disability status | Day to day activities limited a lot/a little | 342,275 (4.42) | 45 (6.34) | 1,870 (40.17) |
|  | Day to day activities not limited | 7,405,070 (95.58) | 665 (93.66) | 2,785(59.83) |
| Highest level of qualifications of the household reference person | No qualifications | 1,098,950 (14.18) | 80 (11.35) | 970 (20.84) |
|  | Below degree qualifications | 4,193,145 (54.12) | 370 (52.48) | 2,505 (53.81) |
|  | Degree level or above | 2,399,980 (30.98) | 255 (36.17) | 1,115 (23.95) |
| NS-SEC of the household reference person | Higher managerial, administrative and professional occupations | 2,058,840 (26.57) | 210 (29.58) | 1,005 (21.61) |
|  | Intermediate occupations | 1,802,865 (23.27) | 155 (21.83) | 1,070 (23.01) |
|  | Routine and manual occupations | 2,413,730 (31.16) | 190 (26.76) | 1,590 (34.19) |
|  | Never worked and long term unemployed | 436,980 (5.64) | 50 (7.04) | 445 (9.57) |
| Main Language | Main language is English | 7,352,740 (94.91) | 685 (96.48) | 4,380 (94.09) |
|  | Main language is not English | 394,605 (5.09) | 25 (3.52) | 275 (5.91) |
| Religion of the household reference person | Christian | 4,233,740 (54.65) | 375 (53.96) | 2,330 (50.11) |
|  | Muslim | 630,550 (8.14) | 25 (3.60) | 565 (12.15) |
|  | No religion | 2,151,475, (27.77) | 240 (34.53) | 1,320 (28.39) |
|  | Other religion | 290,530 (3.75) | 25 (3.60) | 145 (3.12) |
| Region of residence | East Midlands | 668,380 (8.63) | 65 (9.15) | 410 (8.81) |
|  | East of England | 879,800 (11.36) | 70 (9.86) | 485 (10.42) |
|  | London | 1,135,615 (14.66) | 80 (11.27) | 660 (14.18) |
|  | North East | 365,060 (4.71) | 35 (4.93) | 255 (5.48) |
|  | North West | 1,033,750 (13.34) | 125 (17.61) | 730 (15.68) |
|  | South East | 1,295,605 (16.72) | 110 (15.49) | 675 (14.50) |
|  | South West | 752,355 (9.71) | 90 (12.68) | 385 (8.27) |
|  | West Midlands | 847,670 (10.94) | 60 (8.45) | 530 (11.39) |
|  | Yorkshire and The Humber | 769,110 (9.93) | 75 (10.56) | 525 (11.28) |
| Sex | Male | 3,957,370 (51.08) | 450 (63.38) | 2,790 (59.94) |
|  | Female | 3,789,975 (48.92) | 260 (36.62) | 1,865 (40.06) |

Notes for Table 1:
1. For disclosure control purposes, counts are not shown where there are fewer than 10 deaths in a group; this is noted by the shorthand symbol [c]. 2. Characteristics derived for an individual's household reference person contain "Unknown" and "Not stated" groups where a response is not available. Where responses are "Unknown" or "Not stated" results have not been reported, percentages within these groups may not sum to 100.

### Rates of suicide and non-suicide deaths per 100,000 people

Rates of suicide were higher in males (2.09 per 100,000 person-years, 95% CI 1.90-2.88) compared with females (1.27 per 100,000 person-years, 95% CI 1.11-1.42). For ethnic group, we found the highest rate of suicide among CYP from the Mixed or Multiple ethnic group (2.51 per 100,000 person-years, 95% CI 1.85-3.33) (Table 2). Rates of suicide increased with age, with the highest rate in those aged 17 years (4.37 per 100,000 person-years, 95% CI 3.86-4.88). Rates of suicide were also highest in CYP: with a disability; whose main language was English; living in the North and South West of England; where the HRP’s highest level of education was degree-level or above; and where the HRP was long-term unemployed or had never worked.

**Table 2.**
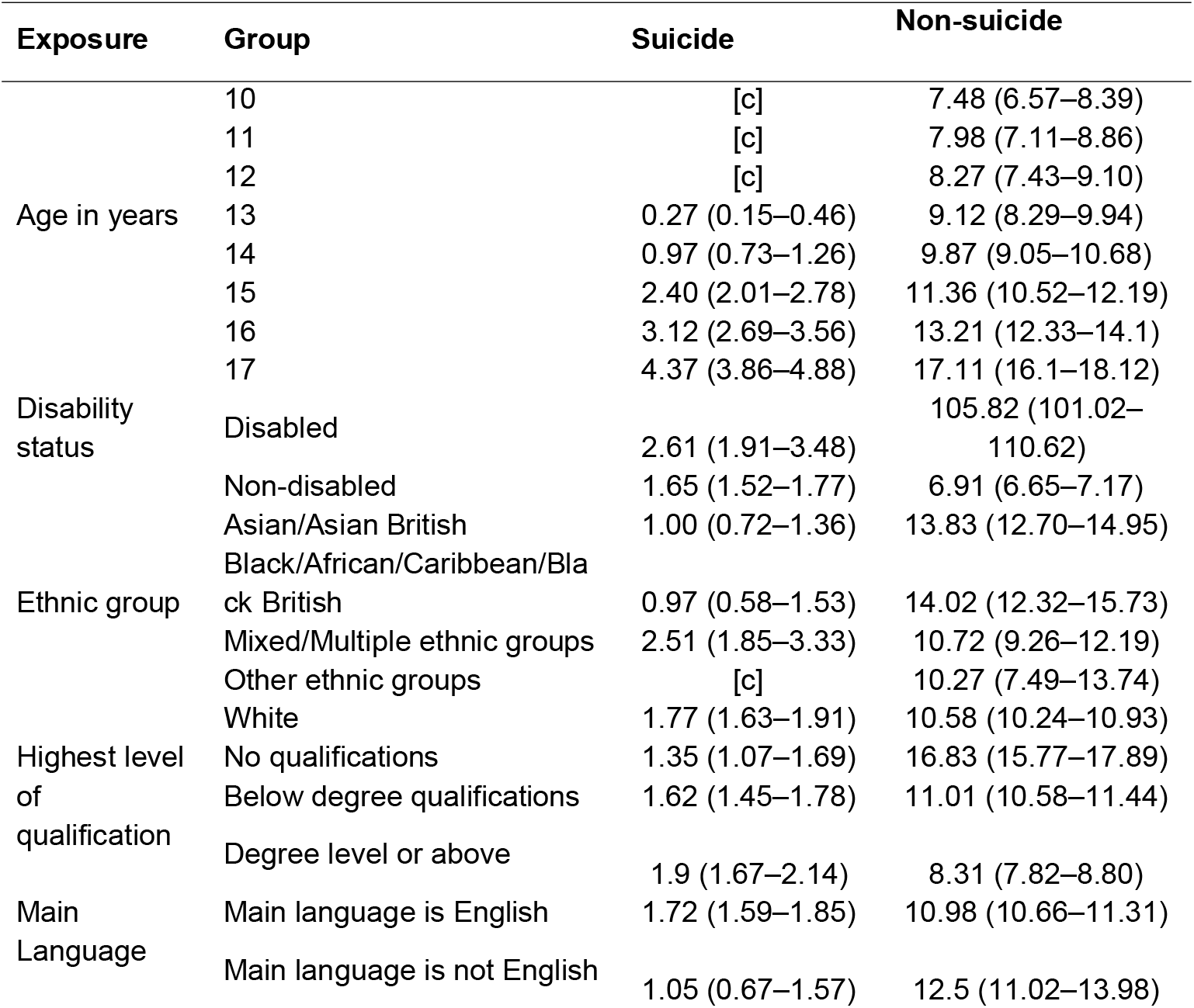
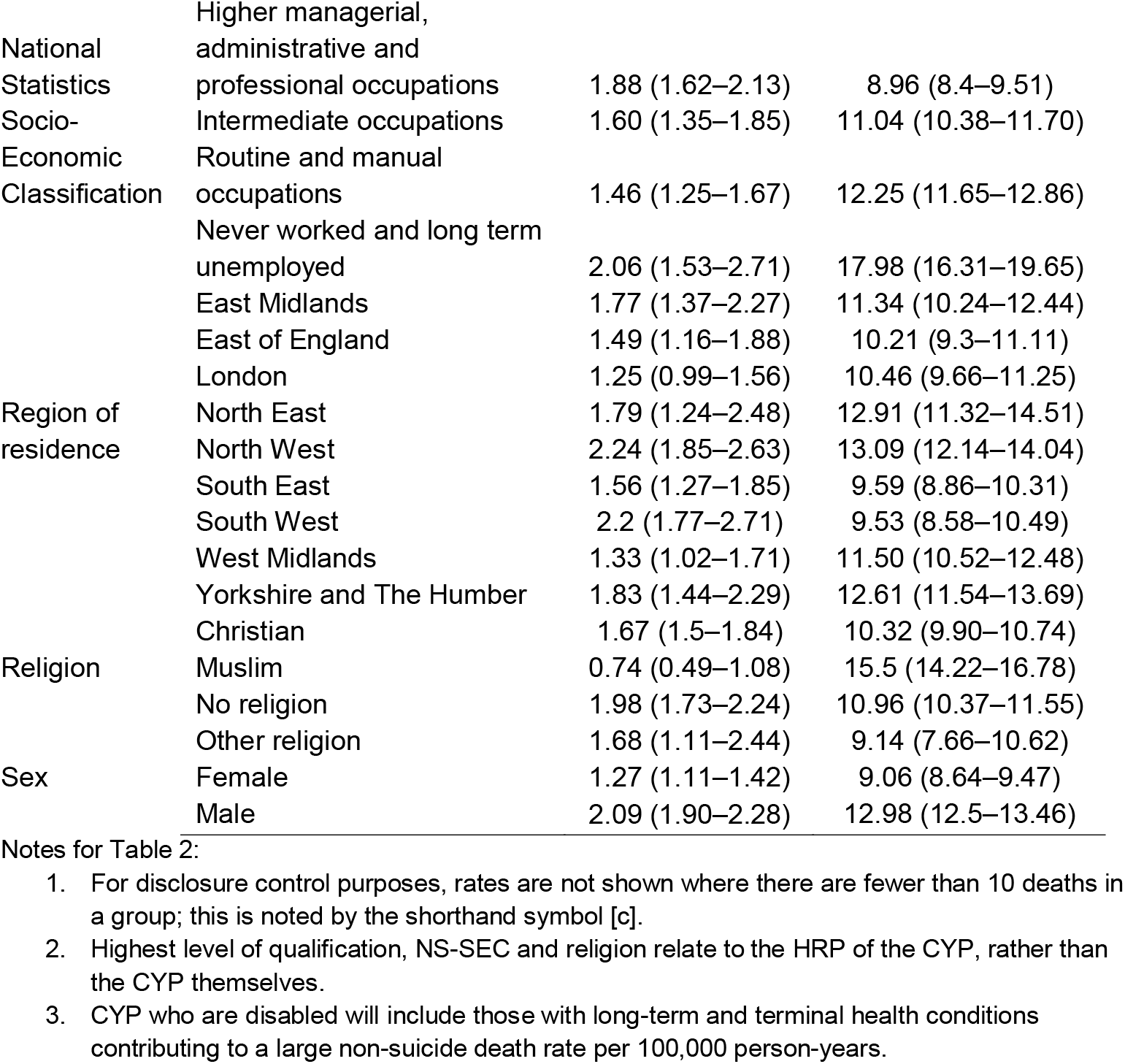
Rates of suicide and other deaths per 100,000 person-years (95% CI) by socio-demographic characteristics.

For age, sex, disability and region we see similar rates for non-suicide deaths. However, for other socio-demographic characteristics we see different patterns emerging. We see a greater risk for non-suicide deaths in those who are Asian/Asian British, Black/African/Caribbean/Black British whilst these carry the lowest rate of suicide death in CYP. Additionally, CYP where the HRP is Muslim has an increased risk of non-suicide death but a lower risk of suicide death. We see the opposite pattern of risk for non-suicide deaths compared to suicide deaths for the highest level of qualification of their HRP.

### Multivariable analysis

Minimally and further adjusted IRRs for suicide in CYP can be seen in Table 3. The rate of suicide was lower for CYP who were female (IRR = 0.61, 95% CI = 0.52 - 0.71) compared to male; and higher for those who identified as being from a Mixed or Multiple ethnic group (IRR = 1.51, 95% CI = 1.12 – 2.02) compared to those who were White. The Asian/Asian British (IRR = 0.59, 95% CI = 0.43 - 0.81) and Black, African, Caribbean, or Black British (IRR = 0.57, 95% CI = 0.36 - 0.91) ethnic groups had the lowest rate of suicide compared to the White ethnic group. Disabled CYP had a higher rate of suicide (IRR = 1.41; 95% CI = 1.04 – 1.90) compared to non-disabled CYP. When adjusting for other characteristics besides age and sex, the pattern of risk remained the same for sex, ethnic group and disability status.

**Table 3.**
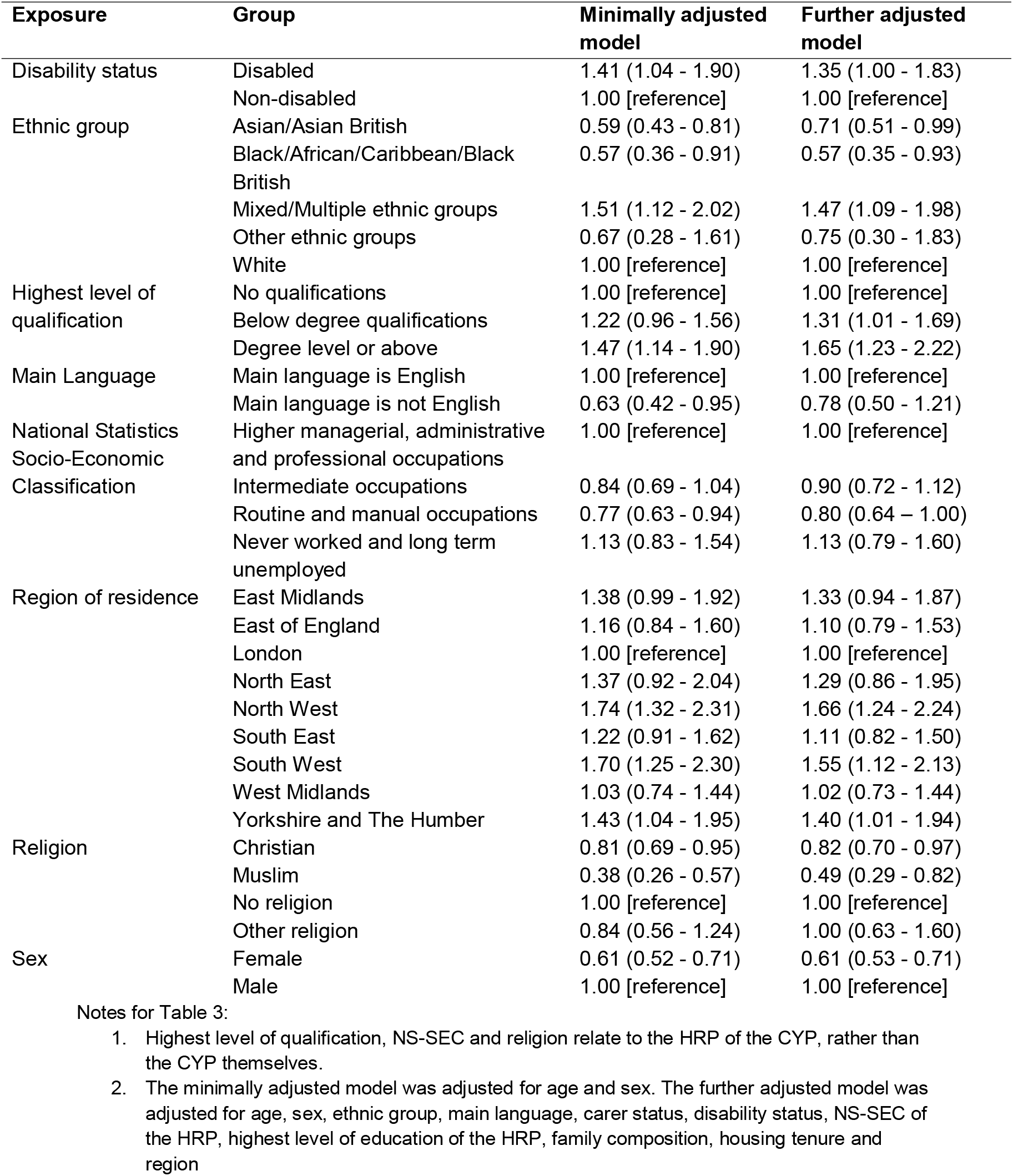
Incidence rate ratios (95% CI) for suicide by socio-demographic characteristics.

The rate of suicide was lower for CYP whose HRP was in a routine or a manual occupation (IRR = 0.77; 95% CI = 0.63 – 0.94) compared to where the HRP was in a higher managerial occupation. Additionally, for CYP whose HRP’s highest level of education was a degree-level or above had the highest rate of suicide (IRR = 1.47; 95% CI = 1.14 – 1.90). When adjusting for other characteristics additionally those whose HRP had below a degree level qualification (IRR = 1.31, 95% CI = 1.01 – 1.69) also had an increased risk of suicide compared to those whose HRP had no formal qualifications. The rate of suicide for CYP was higher than in London in all regions except the West Midlands (IRR = 1.03; 95% CI = 0.74 - 1.44).

## Discussion

We found the risk for suicide was highest in CYP who: were male, were in the Mixed or Multiple ethnic groups, and were disabled. We also found the relative rate of suicide in CYP was higher where the HRP had no religious affiliation and had degree-level qualifications or above. To our knowledge, this study is the first to examine the socio-demographic risk factors for suicide in CYP aged 10 to 17 years old using administrative and census data with near-whole population coverage.

Recent research in England and Wales on socio-demographic inequalities in suicide in adults found elevated risk in White and Mixed/Multiple ethnic groups^14,22^; they also found evidence of sex-specific heterogeneity, with the risk of suicide being higher in these ethnic groups for females but not for males. Our findings further support the increased risk of suicide for CYP in the Mixed/Multiple ethnic group, who were 1.5 times more likely to die by suicide than those who were White, with the Asian and Black ethnic groups. This provides evidence that the ethnic disparities in suicide present in the adult population also are also found in CYP.

The rate of suicide was higher in disabled CYP. This group being at a greater risk aligns with previous research on adults^14^, and in research published in those aged 6 years old and above diagnosed with severe physical conditions^23^. Additionally, our results for age and sex also aligns with existing evidence showing an increased risk of suicide in males and in older CYP^3^.

Previous findings have concluded that unemployed adults are at greater risk of suicide^14,24^ as well individuals in lower skilled occupations^25^. Financial adversity and unemployment have been identified as key risk factors for suicide, in particular in middle-aged men^4^.

However, in CYP, we found an increase of suicide for CYP where the NS-SEC of the HRP is in a routine or a manual occupation. A CYP’s socio-economic position will likely be determined by their caregivers and other household factors. This suggests a less clear link between SES and the risk of suicide in CYP with our results not finding a high SES to be a protective factor. Socio-economic position during childhood is likely to have an impact on an individual (particularly adult health) over a life course^26^, which may indicate why similar studies on adults have shown a linear pattern with the risk of suicide increasing as the NS-SEC class levels decreased^14^.

Our findings show that CYP living in households where the HRP had higher levels of qualifications experienced the highest rates of suicide. A meta-analysis comparing parental education and the risk of suicidal behaviours in youths showed varying patterns of risk, with some studies exhibiting higher risk in households where parents were more educated^15^. however, the meta-analysis concluded there was a small but positive association between lower parental education and risk of suicidal ideation in youths^15^. Within the same study, a moderator analysis on geographical region and found that studies from Eastern and Southern Asia had increased risk of youth suicide attempts when parents’ education was higher^15^. Our study, to our knowledge, is the first population-level evidence of a link between higher levels of parental education and a increased risk of suicide in CYP in England.

The relationship between the education level of a CYP’s household and risk of suicide may be mediated by other factors that we were unable to observe in the current study such as parents or caregiver working in high-pressure jobs or CYP feeling the need to live up to high academic expectations. Research has identified positive associations between academic pressures and self-reported depression and anxiety^27^, and other research found nearly a third (32%) of CYP who died by suicide between 2014 and 2016 had been experiencing academic pressures^28^. Other familial factors such as low parental monitoring have been indicated in research as having a significantly increased risk of suicidal ideation, suicidal attempts and self-injury in young children (aged 9 to 10 years) in the US^10^.

A meta-analysis into religion and suicide concluded that it is widely accepted that religion can be a protective factor for suicide, although there are aspects which may increase suicide risk^29^. Analysis in the US on suicidal behaviours in adolescents found minimal differences between those with regular religious activity participation and those with little or no religious participation, however there was a positive interaction effect for females in further analyses^30^. In our analysis, we found a significantly reduced risk of suicide in CYP whose HRP was Christian or Muslim, compared to those with no religious affiliation, coherent with research suggesting religion is protective. It is important to note that in our study, census returns are likely to have been completed by a parent or guardian. Research suggests that adolescence involvement in religion is likely to be influenced by parental involvement or family norms^30,31^.

### Strengths and limitations

This study is the first to our knowledge to use a large, population-level linked dataset to understand socio-demographic risk factors for suicide in CYP aged 10 to 17 years in England. Using the 2011 Census allowed us to investigate the demographic characteristics of CYP and socio-economic factors related to the households in which they resided.

Due to the low number of suicides in CYP, we were unable to stratify the analysis by sex to investigate risk factors for males and females separately. With growing rates of suicides in female CYP in recent years^3^, it will be important to investigate this group in the future when more data is available, to understand specific risks in this group.

The socio-demographic characteristics in this study are taken from a snapshot in time at the 2011 Census. Our cohort was followed until the end of December 2022, so time-varying characteristics such as main language, household NS-SEC could have changed over this period. This introduces measurement error into our exposures and covariates. The HRP highest level of qualifications are less likely to change compared to a characteristic like NS-SEC.

We controlled for some health and disability factors recorded in the 2011 Census data however we did not control specific health conditions or mediating factors such as the child’s educational attainment and school absenteeism^32^, which may impact mental health and the overall risk of suicide. Research has identified that a history of self-harm and psychiatric treatment increases the risk of suicide in children and adolescents^12^, controlling for this in a model with an outcome of suicide would be misguided. Future research could incorporate additional factors and the impact on suicide risk. However, for this analysis, we cannot rule out residual confounding factors and therefore our estimates should not be interpretated as causal relationships.

There will be some groups who are potentially vulnerable to suicide but are not captured in this analysis, such as those not enumerated on the 2011 Census, including recent migrants. and Census respondents who we were unable to link to the 2011-2013 NHS Patient Registers (for example, due to not being registered with a general practitioner).

### Conclusions

CYP belonging to certain demographic risks, for example those who are male, disabled and belong to Mixed or Multiple ethnic groups, experience elevated rates of suicide. Moreover, the highest rates of suicide are found in CYP with well-educated parents – contrary to the educational gradient generally observed for most other adverse health outcomes in the general population. Therefore, whilst our findings support some similar risk factors in CYP as have been found for adults, the relationship between measures of SES and risk of suicide appears to be more complex in CYP. Further research is needed to understand the underlying mechanism driving these different risk factors for suicide in CYP, so that healthcare policymakers and providers may consider this evidence base when formulating future public health strategies aimed at reducing suicide risk in CYP.

## Data availability

The source data are not publicly available and are subject to controlled access due to their sensitive nature. Census 2011 and death registration data are available through the Integrated Data Service (IDS). Details of the application requirements and process, and the use of data, are available at https://integrateddataservice.gov.uk/how-to-access-the-integrated-data-service.

## Code availability

The code for the project is available at https://github.com/ONS-Health-modelling-hub/risk_factors_suicide_cyp

## Author contributions

ES and IW conceived, acquired funding and designed the study. RM led the data analysis with support from; EJ, ES and IW. RM and EJ undertook quality assurance of the code and data. ES, IW, RM, EJ, DA and VN quality assured the outputs and the interpretation of them. ES, IW, RM, EJ, CR, DA and VN contributed to critical revision of the manuscript and approved the final manuscript.

## Acknowledgements

The authors would like to thank David Mais (Office for National Statistics) for his contributions to reviewing the analysis and in the Patient and Public Involvement and Engagement.

We would like to thank the participants of our two public and patient advisory groups. Our groups comprised of young adults with experiences of suicidality or supporting others with suicidality, and secondly adults who had lost a child or family member to suicide, or who were supporting children or family members with suicidality either in a personal or professional capacity. Members of our public and patient advisory groups reviewed the results of the analysis and provided feedback. We would like to thank Papyrus UK and The Mix for facilitating these workshops. □□

We would also like to thank members of the Mutual Support for Mental Health Research (MS4MH-R) Patient and Public Involvement and Engagement group affiliated with the NIHR Greater Manchester Patient Safety Research Collaboration and Centre for Mental Health and Safety at The University of Manchester□for their contribution to the interpretation of results and reviewing of publications.□

## References

1. Office for National Statistics (ONS). Deaths registered in England and Wales: 2023 [Internet]. 2024 [cited 2024 Dec 20]. Available from: https://www.ons.gov.uk/peoplepopulationandcommunity/birthsdeathsandmarriages/deaths/bulletins/deathsregistrationsummarytables/2023

2. World Health Organization. Suicide [Internet]. 2023 [cited 2024 Jun 27]. Available from: https://www.who.int/news-room/fact-sheets/detail/suicide

3. Office for National Statistics. Suicides in England and Wales: 2023 registrations [Internet]. 2024 [cited 2024 Nov 20]. Available from: https://www.ons.gov.uk/peoplepopulationandcommunity/birthsdeathsandmarriages/deaths/bulletins/suicidesintheunitedkingdom/2023

4. Department of Health and Social Care. Suicide prevention strategy for England: 2023 to 2028 [Internet]. 2023 [cited 2024 Jun 27]. Available from: https://www.gov.uk/government/publications/suicide-prevention-strategy-for-england-2023-to-2028

5. NHS England. NHS Talking Therapies, for anxiety and depression, Annual reports, 2023-24 [Internet]. 2024 [cited 2025 Feb 12]. Available from: https://digital.nhs.uk/data-and-information/publications/statistical/nhs-talking-therapies-for-anxiety-and-depression-annual-reports/2023-24

6. Childrens Commissioner. Children’s Mental Health Services 2021-22 [Internet]. 2023. Available from: https://assets.childrenscommissioner.gov.uk/wpuploads/2023/03/Childrens-Mental-Health-Services-2021-2022-1.pdf

7. Turecki G, Brent DA, Gunnell D, O’Connor RC, Oquendo MA, Pirkis J, et al. Suicide and suicide risk. Nat Rev Dis Prim. 2019 Oct 24;5(1):74.

8. Suicide by Children and Young People National Confidential Inquiry into Suicide and Homicide by People with Mental Illness [Internet]. 2017. Available from: www.hqip.org.uk/national-programmes/a-z-

9. Cybulski L, Ashcroft DM, Carr MJ, Garg S, Chew-Graham CA, Kapur N, et al. Risk factors for nonfatal self-harm and suicide among adolescents: two nested case– control studies conducted in the UK Clinical Practice Research Datalink. J Child Psychol Psychiatry Allied Discip. 2021;

10. Deville DC, Whalen D, Breslin FJ, Morris AS, Khalsa SS, Paulus MP, et al. Prevalence and Family-Related Factors Associated with Suicidal Ideation, Suicide Attempts, and Self-injury in Children Aged 9 to 10 Years. JAMA Netw Open. 2020;3(2).

11. Ruch DA, Heck KM, Sheftall AH, Fontanella CA, Stevens J, Zhu M, et al. Characteristics and precipitating circumstances of suicide among children aged 5 to 11 years in the united states, 2013-2017. JAMA Netw Open. 2021 Jul 27;4(7):E2115683.

12. Hawton K, Bergen H, Kapur N, Cooper J, Steeg S, Ness J, et al. Repetition of self-harm and suicide following self-harm in children and adolescents: Findings from the Multicentre Study of Self-harm in England. J Child Psychol Psychiatry Allied Discip. 2012 Dec;53(12):1212–9.

13. Lorant V, Kunst AE, Huisman M, Costa G, Mackenbach J. Socio-economic inequalities in suicide: A European comparative study. Br J Psychiatry. 2005 Jul;187(JULY):49–54.

14. Ward IL, Finning K, Ayoubkhani D, Hendry K, Sharland E, Appleby L, et al. Sociodemographic inequalities of suicide: a population-based cohort study of adults in England and Wales 2011-21. Eur J Public Health. 2024 Apr 1;34(2):211–7.

15. Chen PJ, MacKes N, Sacchi C, Lawrence AJ, Ma X, Pollard R, et al. Parental education and youth suicidal behaviours: a systematic review and meta-analysis. Epidemiol Psychiatr Sci. 2022;31(e19):1–16.

16. Nafilyan V, Bosworth M, Morgan J, Ayoubkhani D, Dolby T, Groom P, et al. Cohort Profile: The Public Health Data Asset, 2011 cohort. Int J Epidemiol [Internet]. 2024 Feb 1;53(1):dyad194. Available from: 10.1093/ije/dyad194

17. Office for National Statistics. Impact of registration delays on mortality statistics in England and Wales: 2022 [Internet]. 2024 [cited 2024 Nov 20]. Available from: https://www.ons.gov.uk/peoplepopulationandcommunity/birthsdeathsandmarriages/deaths/articles/impactofregistrationdelaysonmortalitystatisticsinenglandandwales/latest

18. Office for National Statistics (ONS). Suicide rates in the UK QMI [Internet]. 2019 [cited 2025 Mar 7]. Available from: https://www.ons.gov.uk/peoplepopulationandcommunity/birthsdeathsandmarriages/deaths/methodologies/suicideratesintheukqmi

19. Office for National Statistics. Suicides in the UK: 2014 registrations [Internet]. 2016 [cited 2025 Jan 24]. Available from: https://www.ons.gov.uk/peoplepopulationandcommunity/birthsdeathsandmarriages/deaths/bulletins/suicidesintheunitedkingdom/2014registrations#methodological-changes

20. Adelstein A, Mardon C. Suicides 1971-74. Popul Trends. 1975;2:13–8.

21. Office for National Statistics. The National Statistics Socio-economic classification (NS-SEC) [Internet]. [cited 2025 Jan 16]. Available from: https://www.ons.gov.uk/methodology/classificationsandstandards/otherclassifications/thenationalstatisticssocioeconomicclassificationnssecrebasedonsoc2010

22. Knipe D, Moran P, Howe LD, Karlsen S, Kapur N, Revie L, et al. Ethnicity and suicide in England and Wales: a national linked cohort study. The Lancet Psychiatry. 2024 Aug;11(8):611–9.

23. Nafilyan V, Morgan J, Mais D, Sleeman KE, Butt A, Ward I, et al. Risk of suicide after diagnosis of severe physical health conditions: A retrospective cohort study of 47 million people. Lancet Reg Heal - Eur. 2023;25.

24. Berkelmans G, van der Mei R, Bhulai S, Gilissen R. Identifying socio-demographic risk factors for suicide using data on an individual level. BMC Public Health. 2021 Dec 1;21(1702).

25. Milner A, Spittal MJ, Pirkis J, LaMontagne AD. Suicide by occupation: Systematic review and meta-analysis. Br J Psychiatry. 2013 Dec 2;203(6):409–16.

26. Arpino B, Gumà J, Julià A. Early-life conditions and health at older ages: The mediating role of educational attainment, family and employment trajectories. PLoS One. 2018;13(4):1–17.

27. Steare T, Gutiérrez Muñoz C, Sullivan A, Lewis G. The association between academic pressure and adolescent mental health problems: A systematic review. J Affect Disord. 2023;339(June):302–17.

28. Rodway C, Tham SG, Ibrahim S, Turnbull P, Kapur N, Appleby L. Children and young people who die by suicide: childhood-related antecedents, gender differences and service contact. BJPsych Open. 2020;6(3):1–9.

29. Gearing RE, Alonzo D. Religion and Suicide: New Findings. J Relig Health [Internet]. 2018;57(6):2478–99. Available from: 10.1007/s10943-018-0629-8

30. Nkansah-Amankra S, Diedhiou A, Agbanu SK, Agbanu HLK, Opoku-Adomako NS, Twumasi-Ankrah P. A longitudinal evaluation of religiosity and psychosocial determinants of suicidal behaviors among a population-based sample in the United States. J Affect Disord. 2012;139(1):40–51.

31. Lawrence RE, Oquendo MA, Stanley B. Religion and Suicide Risk: A Systematic Review. Arch Suicide Res. 2016 Jan 2;20(1):1–21.

32. Epstein S, Roberts E, Sedgwick R, Polling C, Finning K, Ford T, et al. School absenteeism as a risk factor for self-harm and suicidal ideation in children and adolescents: a systematic review and meta-analysis. Vol. 29, European Child and Adolescent Psychiatry. Springer Science and Business Media Deutschland GmbH; 2020. p. 1175–94.

