## Supplementary tables for "Socio-demographic differences in the risk of suicides in children and young people: a population level linked study in England, 2011 to 2022"

### **Supplementary Table 1: All covariates and groupings. All variables are defined from 2011 Census data**

| **Variable** | **Levels** |
| --- | --- |
| Age | Continuous spline in years calculated on Census day |
| Disability status | Day to day activities limited a lot/a little,  Day to day activities not limited |
| Ethnicity | Asian/Asian British, Black/African/Caribbean/Black British, Mixed/multiple ethnic groups, Other ethnic group, White |
| Highest level of qualifications of the household reference person | No qualifications, Below degree level qualifications, Degree level or above |
| Main Language | Main language is English, Main language is not English |
| National Statistics Socio-economic classification (NS-SEC) of the household reference person | Higher managerial, administrative and professional occupations, Intermediate occupations, Routine and manual occupations, Never worked and long term unemployed |
| Region of residence | East Midlands, East of England, London, North East, North West, South East, South West, West Midlands, Yorkshire and The Humber |
| Religion of the household reference person | Christian, Muslim, No religion, Other religion |
| Sex | Male, Female |

### **Supplementary table 2: Intentional self-harm ICD-10 codes (X60-X84) from Chapter XX: External causes of morbidity and mortality**

| **ICD-10 code** | **ICD-10 definition** |
| --- | --- |
| X60 | Intentional self-poisoning by and exposure to nonopioid analgesics, antipyretics and antirheumatics |
| X61 | Intentional self-poisoning by and exposure to antiepileptic, sedative-hypnotic, antiparkinsonism and psychotropic drugs, not elsewhere classified |
| X62 | Intentional self-poisoning by and exposure to narcotics and psychodysleptics [hallucinogens], not elsewhere classified |
| X63 | Intentional self-poisoning by and exposure to other drugs acting on the autonomic nervous system |
| X64 | Intentional self-poisoning by and exposure to other and unspecified drugs, medicaments and biological substances |
| X65 | Intentional self-poisoning by and exposure to alcohol |
| X66 | Intentional self-poisoning by and exposure to organic solvents and halogenated hydrocarbons and their vapours |
| X67 | Intentional self-poisoning by and exposure to carbon monoxide and other gases and vapours |
| X68 | Intentional self-poisoning by and exposure to pesticides |
| X69 | Intentional self-poisoning by and exposure to other and unspecified chemicals and noxious substances |
| X70 | Intentional self-harm by hanging, strangulation and suffocation |
| X71 | Intentional self-harm by drowning and submersion |
| X72 | Intentional self-harm by handgun discharge |
| X73 | Intentional self-harm by rifle, shotgun and larger firearm discharge |
| X74 | Intentional self-harm by other and unspecified firearm discharge |
| X75 | Intentional self-harm by explosive material |
| X76 | Intentional self-harm by smoke, fire and flames |
| X77 | Intentional self-harm by steam, hot vapours and hot objects |
| X78 | Intentional self-harm by sharp object |
| X79 | Intentional self-harm by blunt object |
| X80 | Intentional self-harm by jumping from a high place |
| X81 | Intentional self-harm by jumping or lying before moving object |
| X82 | Intentional self-harm by crashing of motor vehicle |
| X83 | Intentional self-harm by other specified means |
| X84 | Intentional self-harm by unspecified means |

### **Supplementary table 3: Event of undetermined intent ICD-10 codes (Y10-Y34) from Chapter XX: External causes of morbidity and mortality**

| **ICD-10 code** | **ICD-10 definition** |
| --- | --- |
| Y10 | Poisoning by and exposure to nonopioid analgesics, antipyretics and antirheumatics, undetermined intent |
| Y11 | Poisoning by and exposure to antiepileptic, sedative-hypnotic, antiparkinsonism and psychotropic drugs, not elsewhere classified, undetermined intent |
| Y12 | Poisoning by and exposure to narcotics and psychodysleptics [hallucinogens], not elsewhere classified, undetermined intent |
| Y13 | Poisoning by and exposure to other drugs acting on the autonomic nervous system, undetermined intent |
| Y14 | Poisoning by and exposure to other and unspecified drugs, medicaments and biological substances, undetermined intent |
| Y15 | Poisoning by and exposure to alcohol, undetermined intent |
| Y16 | Poisoning by and exposure to organic solvents and halogenated hydrocarbons and their vapours, undetermined intent |
| Y17 | Poisoning by and exposure to other gases and vapours, undetermined intent |
| Y18 | Poisoning by and exposure to pesticides, undetermined intent |
| Y19 | Poisoning by and exposure to other and unspecified chemicals and noxious substances, undetermined intent |
| Y20 | Hanging, strangulation and suffocation, undetermined intent |
| Y21 | Drowning and submersion, undetermined intent |
| Y22 | Handgun discharge, undetermined intent |
| Y23 | Rifle, shotgun and larger firearm discharge, undetermined intent |
| Y24 | Other and unspecified firearm discharge, undetermined intent |
| Y25 | Contact with explosive material, undetermined intent |
| Y26 | Exposure to smoke, fire and flames, undetermined intent |
| Y27 | Contact with steam, hot vapours and hot objects, undetermined intent |
| Y28 | Contact with sharp object, undetermined intent |
| Y29 | Contact with blunt object, undetermined intent |
| Y30 | Falling, jumping or pushed from a high place, undetermined intent |
| Y31 | Falling, lying or running before or into moving object, undetermined intent |
| Y32 | Crashing of motor vehicle, undetermined intent |
| Y33 | Other specified events, undetermined intent |
| Y34 | Unspecified event, undetermined intent |

### **Supplementary Table 4: Sample Flow of Population**

| **Stage** | **Count** |
| --- | --- |
| 2011 Census Population | 56,963,310 |
| Sample of 2011 Census respondents able to be linked to 2011-2013 NHS Patient Registers and have a usual residence flag | 50,189,575 |
| Sample who were living in England during the study dates | 47,454,355 |
| Sample of who were alive at the end of the study (31^st^ December 2022) or died between 2011 Census day (27 March 2011) and end of study | 47,454,210 |
| Sample who were aged between 10 and 17 years on 2011 Census day and end of study | 7,747,345 |
